# An observational study identifying highly tuberculosis-exposed, HIV-1-positive but persistently TB, tuberculin and IGRA negative persons with *M. tuberculosis* specific antibodies in Cape Town, South Africa

**DOI:** 10.1101/2020.07.07.20147967

**Authors:** Elouise E. Kroon, Craig J. Kinnear, Marianna Orlova, Stephanie Fischinger, Sally Shin, Sihaam Boolay, Gerhard Walzl, Ashley Jacobs, Robert J. Wilkinson, Galit Alter, Erwin Schurr, Eileen G. Hoal, Marlo Möller

## Abstract

**Background:** *Mycobacterium tuberculosis (Mtb)* infection is inferred from positive results of T-cell immune conversion assays measuring Mtb-specific interferon gamma production or tuberculin skin test (TST) reactivity. Certain exposed individuals do not display T-cell immune conversion in these assays and do not develop TB. Here we report a hitherto unknown form of this phenotype: HIV-1-positive persistently TB, tuberculin and IGRA negative (HITTIN).

**Methods:** A community-based case-control design was used to systematically screen and identify adults living with HIV (HIV+), aged 35-60 years, who met stringent study criteria, and then longitudinally followed up for repeat IGRA and TST testing. Participants had no history of TB despite living in TB hyper-endemic environments in Cape Town, South Africa with a provincial incidence of 681/100,000. *Mtb*-specific antibodies were measured using ELISA and Luminex.

**Findings:** We identified 48/286 (17%) individuals who tested persistently negative for *Mtb*-specific T-cell immunoreactivity (three negative Quantiferon results and one TST = 0mm) over 206±154 days on average. Of these, 97·2% had documented CD4 counts<200 prior to antiretroviral therapy (ART). They had received ART for 7·0±3·0 years with a latest CD4 count of 505·8±191·4 cells/mm^3^. All HITTIN sent for further antibody testing (n=38) displayed Mtb-specific antibody titres.

**Interpretation:** Immune reconstituted HIV+ persons can be persistently non-immunoreactive to *Mtb*, yet develop species-specific antibody responses. Exposure is evidenced by *Mtb*-specific antibody titres. Our identification of HIV+ individuals displaying a persisting lack of response to TST and IGRA T-cell immune conversion paves the way for future studies to investigate this phenotype in the context of HIV-infection that so far have received only scant attention.

**Funding:** Funding provided by National Institutes of Health [1R01AI124349-01].

**Research in Context section:** *Evidence before this study:* The majority of individuals who are exposed to *Mycobacterium tuberculosis (Mtb)* appear to have natural immunity against developing tuberculosis (TB). A subset of these individuals may clear or control *Mtb* without developing a classical T-cell immune response as measured by a tuberculin skin test (TST) or IGRA. While the gold standard for TB diagnosis is culturing *Mtb* from a specimen, there is no direct test to prove current *Mtb* infection. Hence, infection needs to be inferred from tests that measure *Mtb* T-cell immunoreactivity. Once *Mtb* is inhaled, pulmonary innate immune cells, so-called alveolar macrophages, are the first to make contact with the bacilli. *Mtb*-infected alveolar macrophages traverse from the alveoli to the lung interstitium where *Mtb* is transferred to inflammatory macrophages that present the bacilli to T-cells and initiate the adaptive immune response, including the generation of memory T-cells. The current *Mtb* T-cell immunoreactivity tests indicate infection if positive. A major caveat with these tests is an inability to distinguish between a current infection or a persistent immune response after a previous infection that was cleared. In addition it is important to clarify that persisting negative immunoreactivity does not simply imply that individuals are not infected. They could have had previous immunoreactivity which has reverted, could not have been exposed to *Mtb* or they were exposed but cleared *Mtb* without a classical IFN-γ T-cell immunoreactive response. The latter is a novel concept and previous studies in HIV-negative subjects have focused on this resistance to ‘infection’- or to be more precise failure of IFN-γ T-cell and TST immune conversion to *Mtb*. However, these studies found *Mtb*-specific antibodies in their HIV-negative ‘resister’ subjects. The presence of especially IgG antibodies confirmed and indicated long term exposure to *Mtb* and pointed to current or cleared infection. T-cell responses are required for antigen-specific B-cells to release class-switched IgG. Since these individuals test persistently negative for TST and IFN-γ T-cell immunoreactivity, innate and alternative adaptive IFN-γ independent T-cell responses should be investigated. This is a developing field and as research emerges, it is increasingly important to clearly characterize and define the TB resistance phenotype.

*Added value of this study:* HIV-positive (HIV+) persons are at increased risk of infection with *Mtb* and rapid progression to tuberculosis making the phenotype we describe here especially important in this population segment. However, there is a major lack of data in HIV+ persons describing persisting lack of response to TST and *Mtb* IFN-γ T-cell immune conversion. Here we show that this phenotype can be identified in immune-reconstituted HIV+ persons. We describe the recruitment and define this conversion resistance phenotype as HIV-1-positive persistently TB, tuberculin and IGRA negative (HITTIN). The presence of specific antibodies confirms exposure to *Mtb*. Our study suggests possible innate mechanisms and non-classical adaptive mechanisms that subvert early stages of tuberculosis pathogenesis in a substantial proportion of HIV-infected patients. Harnessing such TST and IFN-γ T-cell independent mechanisms of resistance is of particular interest for prevention of tuberculosis in the HIV+ population.

*Implications of all the available evidence:* Understanding the mechanisms of resistance will enable us to develop TB prevention and treatment modalities.

## Introduction

Classically, tuberculosis (TB) pathogenesis is broadly divided into what is termed latent TB infection (LTBI) and active TB, with only a small proportion of LTBI persons likely to advance to disease. *Mycobacterium tuberculosis (Mtb)* infection in the absence of clinical symptoms is inferred from T-cell based assays such as a positive tuberculin skin test (TST) or interferon gamma release assay (IGRA). Recently, increased attention has been given to the first step of TB pathogenesis, namely infection as measured by TST and IFN-γ T-cell immune conversion. [1] Persons who remain LTBI-negative, i.e. who do not convert their negative IGRA or TST status, and do not develop TB, despite known high *Mtb* exposure are generally considered to be also resistant to active TB. These persons have been termed either “resisters” or “early clearers” [2-5] and several immune and genetic factors have been identified that contribute to this phenotype.[6-10]. Simmons et al proposed baseline criteria to define the resister phenotype. These criteria include the requirement that individuals should have demonstrable prolonged and intense exposure to *Mtb*. In addition, resisters should have persistently negative results in IGRA and TST assays with no history of active TB.[1] This case definition for the resister phenotype was not reached by consensus methodology and more studies are needed to fully describe this phenotype. [11] Recently other studies investigating this phenotype have identified the presence of *Mtb*-specific antibodies in their defined so-called ‘resister’ groups. [12] This challenges current dogma defining LTBI and indicates these persons were previously exposed to *Mtb* and have either current or cleared infection. It is clear that the present measures of *Mtb* infection do not detect the full clinical TB disease spectrum. In addition neither IGRA nor TST or current antibody profiles are able to differentiate between current or cleared paucibacillary infection.[12] Persons with negative baseline TST and IGRA and who do not convert have lower TB incidence rates than those who convert.[13,14] Further research and long term follow-up is essential to understand these emerging and unique innate and adaptive immune profiles in these individuals.

The study of TB resistance is of particular importance in the context of people living with HIV infection in areas of high *Mtb* transmission such as sub-Saharan Africa where nearly 80% of all people suffering from HIV-associated TB reside. People living with HIV are at increased risk of TB disease and molecular epidemiological studies show that HIV-associated TB is usually the result of recent infection and rapid progression to disease, and not reactivation of latent infection. [15-17] The Western Cape, Republic of South Africa (RSA), has one of the highest TB incidences worldwide, and studies have shown that *Mtb* transmission occurs mainly in the community as opposed to the household. [18] In addition, the prevalence of HIV infection is estimated at 12·6% of the general population leading to a substantial number of HIV and *Mtb* co-infections.[19] By employing a community-based study design our objective was to identify a group of HIV-1-positive (HIV+) persons who had undergone a period of very low CD4+ T-cell counts and displayed persistent TST and IGRA negativity in absence of any TB history. In particular we aimed to recruit older persons who, because of their age, would have had prolonged exposure to *Mtb* and despite previous severe immune suppression have never had TB. We hypothesised that we could identify distinct phenotypes to study *Mtb* infection in HIV+ individuals. The use of this cohort is to facilitate the study of mechanisms of resistance to TB among HIV+ individuals, including genetic and immunological studies of the phenotype. Focusing on HIV+ patients who experienced a period of low CD4 T cells counts and who remained TB free despite living in settings of high *Mtb* transmission provided a background filter for possible TB resistance. Upon recruitment, however, participants had to be on longterm ART since this reconstitutes IFN-γ CD4 T-cell responses necessary for TST and IGRA assays. We defined these persons as HIV-1-positive persistently TB, tuberculin and IGRA negative (HITTIN). We estimated the frequency of the HITTIN phenotype to be approximately 17% of the enrolled group of HIV+ persons (48/286). Our results demonstrated a distinct phenotype that can be expressed by people living with HIV infection even after they underwent a period of very low CD4+ counts.

## Methods

### Ethics

The study was approved by the Health Research Ethics Committee of Stellenbosch University (N16/03/033 and N16/03/033A) and the Faculty of Health Sciences Human Research Ethics Committee of the University of Cape Town (755/2016 and 702/2017). Ethics approval for the active TB study participants was obtained from the Faculty of Health Sciences Human Research Ethics Committee of the University of Cape Town (012/2007). Additional approval was obtained from the City of Cape Town and Western Cape government for access to the relevant clinics.

### Study setting

The Western Cape province of South Africa has a very high *Mtb* transmission with an annual rate of TST conversion estimated at 7.9% among young adolescents.[20] In the age range 31-35 years, 80-90% of the population display TST reactions ≥ 10mm consistent with reported community prevalence of 69-80%.[20,21] Using age (35-60 years old) as a surrogate for exposure frequency we screened older persons in a high TB incidence environment to identify our phenotype of interest. In 2018, the TB incidence in South Africa was 520 per 100,000 across all age groups [22] and the latest available TB incidence (2015) in the Western Cape was 681 per 100,000 across all age groups.[23] The HIV prevalence in the Western Cape is 12.6%.[18,24] Among HIV-infected persons the cumulative TB risk is greatly increased during periods of low CD4 counts both before and during antiretroviral therapy (ART).[25-27] For ART naïve HIV-infected persons with CD4 counts <100 cells/mm^3^ TB incidence in the recent past reached as high as 30 cases/100 person-years.[25-27]

In the enrolment area, HIV+ patients visit community healthcare clinics (CHCs) every two to six months to access ART. The study focussed on the clinics in the northern suburbs and Khayelitsha which have their tertiary referral centre at Tygerberg Hospital. Tygerberg Hospital is the academic tertiary hospital affiliated with Stellenbosch University. Five out of 10 possible CHCs were included in the study, as these clinics are all within 20km of the lab at Stellenbosch University Faculty of Medicine and Health Sciences and had ART clubs. The daily burden on clinics is extremely high. Once a HIV+ patient is immune reconstituted and ART adherent, they are moved to ART clubs. Adherence club membership is restricted to patients who have been compliant for at least 1 year of ART, with undetectable viral loads and are otherwise healthy. We focussed our recruitment process on ART club members. All participants including previous TB patients were recruited from these ART clubs. In a pilot study, records of HIV+ persons enrolled in ART adherence clubs were pre-screened during November 2016 to January 2017.

### Identification of population of interest in pilot study

Screening of paper records as well as electronic database management systems were based on enrolment criteria that aimed to maximize exposure to *Mtb* in the absence of documented clinical TB. As part of a pilot study, we used this prescreening of records to identify possible participants who met the criteria for enrolment. First, we targeted our search to HIV+ patients in the age group of 35 years to 60 years since age is a surrogate for cumulative likelihood of infectious exposure and an established strong risk factor for latent TB infection (LTBI) [20]. Second, all potential study participants needed to have had a prolonged period of very low CD4 counts (two CD4+ < 350 cells/mm^3^ counts at least 6 months apart or a single CD4+ count < 200 cells/mm^3^), which was a common occurrence prior to the implementation of more intensive HIV screening and antiretroviral therapy (ART) in South Africa. Third, at the time of enrolment we identified potential study participants who had been on ART for at least one year and were immune reconstituted with the latest CD4+ > 200 cells/mm^3^ and no history of acquired immune deficiency syndrome (AIDS) defining illnesses in the last year. Fourth, stratification was done, based on clinic record of previous TB.

This pilot study showed that 29% of club members (136/472) fulfilled the CD4 count criteria for potential enrolment in the main study. The same criteria were applied to all participants (Arm 1 and Arm 2) in the main study. Criteria for enrolment for all participants in the pilot and main study were age of 35 to 60 years, HIV+, living in an area of high transmission of *Mtb*, history of living with a low CD4+ count (either with two CD4+ <350 cells/mm^3^ counts at least 6 months apart or a single CD4+ count <200 cells/mm^3^) prior to initiating ART, and immune reconstitution on ART for at least one year at time of enrolment with the last CD4+ count >200 cells/mm^3^.

### Study design

The main study was executed in two parts during February 2017 to May 2019. Part one was a case-control study that recruited participants based on the recruitment criteria established in the pilot study. Participants were prescreened by clinical record review using the same process described in the pilot study and were approached on the day of their regular visit to the community healthcare clinic (CHC) to access ART. Cases were defined as subjects with no documented and reported current or previous TB (Arm 1). Only subjects with a documented history of bacteriologically confirmed TB during ART that had been cured for at least 3 years were included in the control group Arm 2 (Previous TB). During the interview the participant was asked if they had previous TB or not. Previous TB had to be documented and to be shown as previous bacteriologically confirmed TB during ART but cured for the last 3 years. This was found from previous TB treatment files in patient folders. After obtaining written informed consent, each participant was screened for TB using the World Health Organization (WHO) TB symptoms questionnaire (cough of any duration, fever for more than two weeks, unexplained weight loss, drenching night sweats). Any participant suspected of having TB was excluded from the study and referred for TB workup at the local clinic. Other relevant medical history obtained included known medical comorbidities, medication use, relevant HIV history, Isoniazid Preventive Therapy (IPT) history and history of Cotrimoxazole preventative therapy.

Pregnancy and previous interventional studies were exclusion criterion. Previous interventional studies refer to any previous clinical trials or studies that required an intervention. Many clinical trials are also conducted at the clinics selected for recruitment and we did not want to include participants who were participating especially in vaccine trials which could have immunomodulatory effects on results. In our cohort of participants, childhood immunization procedures were variable and inconsistently recorded. Bacille Calmette-Guérin (BCG) status was not recorded. It was not possible for us to ascertain BCG status with confidence, as discussed in Rangaka et al. [28] However, BCG vaccination has been a routine part of the national expanded program on immunization (EPI) in South Africa since 1973. No matching was done. All Arm 1 participants had an IGRA using QuantiFERON®-TB Gold Plus (QFT®- Plus) in tube test to determine further classification for Part two.

Part two of the study expanded the case-control study to be nested in a cohort study. This part of the study focused on participants with no history of previous TB (Arm 1). Individuals were selected based on their Arm 1 IGRA results and grouped into either IGRA negative or positive. The IGRA negative group was followed-up for a second IGRA and TST administration. Three days after the TST administration the IGRA was repeated for a third time. This was to check for boosting as a result of the TST [14]. Those subjects that had a TST = 0mm and a third negative IGRA were designated HITTIN. In summary, HITTIN are HIV+ persons who had experienced a period of very low CD4 counts, who had no symptoms or history of previous TB, had three consecutive negative IGRA readings, and a TST = 0mm (Figure 1). The IGRA positive group was also followed-up for a second IGRA and TST administration. This group did not receive a third IGRA. As part of the cohort design we defined this group of HIV+ persons who tested IGRA positive in two consecutive tests (IGRA double+) and displayed a TST ≥ 5mm (Figure 1). We refer to these participants as HIV-1-positive IGRA positive tuberculin positive (HIT).

**Figure 1:**
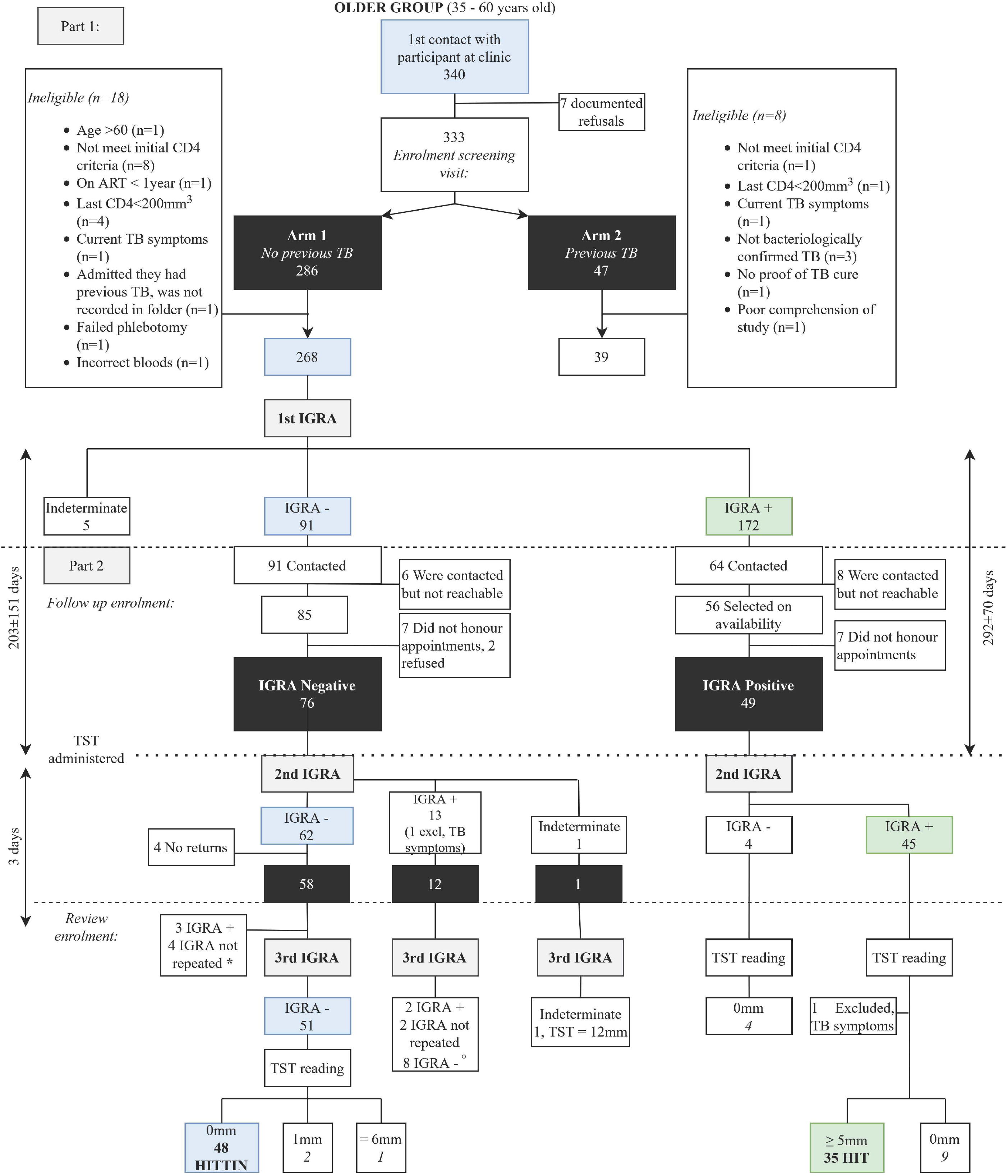
Recruitment flow diagram. The main phenotype derived from **Arm 1 (visit 1)** of the study, was stringently defined according to the criteria discussed under prescreening of records. Eighteen Arm 1 participants were excluded. We recalled as many negative IGRA participants as possible, since this was our phenotype of interest. Fifteen negative IGRA participants were excluded. Sixty-two of the part negative group had a negative second IGRA result. Four participants did not return. *Of the 58 participants who returned for a third visit, three had a positive third IGRA result (2 TST = 0mm, 1 TST = 15mm) and four did not have a third IGRA (TST = 7, 7, 10, 16mm). Fourty-eight participants have three negative IGRA results and a TST reading of 0 mm and were defined as HITTIN (indicated in blue on the flow diagram). °Eight participants tested IGRA negative on the first visit. They had a second positive IGRA result and retested IGRA negative the third time. Six of the eight had TST readings of 0 mm. These six participants likely had false positive IGRA results on the second IGRA test. The HIT group is indicated in green and was selected from 45 persons who tested IGRA positive for a second time and who had TST readings ≥5mm. The timeline on the left reflects the time between the first and the second IGRA of the HITTIN group (203±151 days) and then the time between second to the third IGRA (6±7 days). The timeline on the right reflects the time between the first and the second IGRA of the HIT groups (292±70 days). The TST were read 3 days later with only one read on day 4 and one on day 8. Both of these participants also self-reported persistant lack of TST response.

A subset of samples (n=12) used in the ELISA *Mtb* antibody detection panel was collected from a group of HIV+ persons with microbiologically active TB. [29]

## Immune assays

### IGRA

For IGRAs, participants had 4-5 ml of blood taken and fresh blood was collected for a QuantiFERON®-TB Gold Plus (QFT®-Plus) in tube test (containing contain ESAT-6, CFP-10 and TB 7·7 antigens in two TB antigen tubes) as determined by the supplier. Peptides in the TB1 tubes were selected to elicit CD4+ T-helper lymphocyte response, whereas TB2 has additional peptides for CD8+ cytotoxic T lymphocyte responses. Negative results were interpreted as per the manufacturer’s algorithm [30]. Strict standard operating procedures were adhered to, to minimize operator introduced bias.

### TST

The TST was performed by injecting 0.1 ml of tuberculin purified protein derivative (PPD) intradermally and immediately after drawing blood for QFT®-Plus. This was administered during part 2 of the study when selected Arm 1 participants returned for a second IGRA. For logistical reasons, the initial 104 subjects (49 Arm 1 IGRA positives and 55 Arm 1 IGRA negatives) were tested employing PPD-S (Pasteur-Merieux) while the remaining 20 Arm 1 IGRA negatives subjects were tested with PPD RT23 (Staten Serum Institute). The TST was read on average 72 hours after administration. Medical personnel followed standard operating procedures for each TST administration and reading. A total of eight participants were repeat tested with both PPD-S and RT23 [31]. This could have potentially introduced bias, but the distribution of HITTIN did not differ between Arm 1 IGRA negatives testing with PPD-S (37/55) or PPD RT23 (11/20) (p=0·33, chi-square test).

### *Mtb* antibody detection via ELISA

ELISA plates (Nunc MaxiSorp) were coated with 4μg/ml of a recombinant ESAT-6/CFP-10 fusion protein (Lionex LRP-0074·2, Braunschweig, DE) diluted in carbonate bicarbonate buffer (Sigma Aldrich, St Louis MO) overnight at 4°. Next, plates were washed with PBS containing 0·05% Tween-20 (v/v) and blocked with 3% BSA (Sigma Aldrich) for 1 hour at room temperature (RT). Plates were then washed again and coated with patient serum diluted in blocking buffer for 2 hours before washing. Plates were then coated with a secondary anti-human IgG-Alkaline phosphatase conjugated antibody (Sigma Aldrich A3187) diluted 1:1000 in blocking buffer for 1 hour at RT. After washing, plates were then developed with p-nitrophenyl phosphate (PNPP) (Sigma Aldrich) and optical density (OD) values at 415mm read after 30min incubation. OD of antigen uncoated wells was subtracted from wells coated with antigen at each serum dilution, and values normalized to control serum on each plate.

### *Mtb* antibody detection via Luminex microspheres

Relative concentration of antigen-specific antibody subclass and isotype levels were assessed via a customized Luminex array [32]. Different *Mtb* antigens (PPD, ESAT6/CFP10 and LAM, acquired from BEI resources) and TbAd generously provided by Branch Moody, Brigham and Women’s Hospital) were carboxyl-coupled to fluorescent microspheres (MAGPLEX) (Luminex). Coupling was performed by covalent N-hydroxysuccinimide (NHS)–ester linkages via EDC (Thermo Scientific) and Sulfo-NHS (Thermo Scientific) following the manufacturer’s instructions. 1·2·x10^3^ beads per Luminex region/ antigen were added per well of a 384-well plate (Greiner Bio-one) in Luminex assay buffer containing 0·1% BSA and 0·05% Tween-20. 1:10 in PBS diluted plasma samples were added to each well and incubated for 16 h shaking at 900 rpm at 4°C. The immune complexed beads were then washed three times with 60 μl of Luminex assay buffer using an automated plate washer (Tecan). Phycoerythrin (PE)-coupled IgG1, IgG2-, IgG3-, IgG4, IgA1-, IgA2-, IgM- or bulk IgG-specific detection reagents (Southern Biotech) were added at 1·3 μg/ml and incubated with microspheres for 1 h at room temperature while shaking at 900 rpm. The plate was then washed and read on a flow cytometer (iQue, Intellicyt). Events were gated on the specific bead regions based on bead fluorescence and median fluorescent intensity (MFI) for PE was the readout for each secondary detector and bead region. Samples were run in duplicate per each secondary detection agent.

### Statistical analysis

This is the first report of the study and the choice of sample size was motivated by downstream project aims for WGS and eQTL analysis. The aim was to recruit 60 HIV+ adults (35-60 years) in the HITTIN groups and 30 HIV+ adults (35-60 years) in the HIT group in Arm 1. In Arm 2 we aimed for 20 HIV+ adults (35-60 years) with previous TB. Since more participants tested IGRA positive we reached the target of the HIT group sooner than HITTIN.

Given the previous results in HIV-negative subjects showing that the quality of antibody responses differed between “resisters” and immune converters, we wished to test for such separation in the HIV+ participants. Based on this, previous analysis by Lu et al showed a sample size of 40 for each comparison group was sufficient for antibody LASSO-PLSDA analysis.[12] Hence, our sample size of n=38 (HITTIN) and n=35 (HIT) was sufficient. At the time of shipment for antibody analysis the final 10 HITTIN had not been identified, and were therefore not included in this analysis.

All captured data including laboratory results were quality controlled during the study. Missing data was addressed when it occurred. None of the participants included in the final groups had missing data. Participants who were lost to follow-up were excluded from the study.

For demographic data, statistical analysis was performed using Graphpad Prism Version 8·2·0 for Windows, GraphPad Software, San Diego, California USA, www.graphpad.com. An unpaired t test was used for continuous variables and the Fisher’s exact test for comparing nominal variables. A p-value of <0·05 was considered to be statistically significant.

To compare the HITTIN and HIT groups for isotype data, a Mann-Whitney test was performed. For the correlation analyses, pairwise nonparametric two-tailed spearman correlations for the different subclasses and isotypes against ESAT-6/CFP-10 were performed. P values were corrected for multiple comparisons via Bonferroni correction for 15 tests. (Significance for all analyses were defined as p-value <0·05, *\*p<0*·*05, **p<0*·*01, ***p<0*·*001, ****p<0*·*0001.)* Significance for all analyses were defined as Bonferroni corrected p-value <0·05.

The multivariate analysis was performed as a two-step model on 48 antibody features to assess differences and similarities and their drivers comprehensively between HIT and HITTIN individuals. The PLSDA method was used to decipher which principal components can achieve the maximum separation between groups. PLSDA in comparison to PCA is a supervised method that takes the outcome, so the groupings of HIT and HITTIN into account when calculating the maximum differences, ensuring an optimal separation between the groups. It does this by composing both X and Y variables into a hyperplane and maximizes covariance. A total of 48 features of IgG, IgG1-4, IgA1-2 and IgM against different TB antigens were included in the analysis. Following normalization of the data, a penalty-based least absolute shrinkage and selection operator (LASSO) was applied. LASSO is a way to perform feature reduction and regularization: it downselects the minimum number of features that is able to drive the maximum separation, this makes the model easier to interpret and reduce redundant features that do not add any information. This occurs via penalization of the coefficients of the regression variables. LASSO adds a penalty equal to the absolute value of the magnitude of coefficients, therefore some coefficients can become zero and eliminated resulting in a model with less parameters to separate groups.[33] After feature reduction, Partial least square discriminate analysis (PLSDA) was performed to graph the data and visualize the differences between the groups along the latent variable (LV) 1 and 2. A thousand iterations were applied to the 10-fold nested cross-validation model.

## Results

The study design is depicted in figure 1. Samples were collected over a period of 2.5 years. In total, we obtained informed consent from 340 people, while 7 potential participants declined in part one of the study. After the informed consent, 286 participants were enrolled in Arm 1 (no previous TB). The majority (97.4%) of those enrolled had CD4+ counts of < 200 cells/mm^3^ prior to initiating ART. Of the 286 participants 18 were excluded. Of the 268 remaining participants 91 (34%) tested IGRA negative, 172 (64%) IGRA positive and 5 (2%) were indeterminate. In Arm 2, 47 participants agreed to partake in the study. Seven participants were excluded since they failed the eligibility criteria and one participant was excluded due to lack of comprehension which precluded informed consent.

For the second part of the study participants were selected according to their Arm 1 IGRA result. For the IGRA positive group we recontacted participants and made appointments based on soonest availability. We reached our target number which was sufficient for a balanced design in a reasonable amount of time. Of the sixty participants recontacted, ten did not honour appointments. One participant was excluded prior to the repeat IGRA, after confirming a TB meningitis episode in 2011. One participant was excluded due to presenting with TB symptoms on the 3rd visit when returning for a TST reading. Thirty-five participants have two positive IGRA results and a TST reading of ≥ 5mm. Although the cutoff for TST positive readings was defined as ≥ 5mm, all of the 35 TST readings were ≥ 10mm with a mean reading of 18mm. These 35 participants were defined as HIT.

Seventy-six of the 91 IGRA-negative participants were willing to continue in the study for a second IGRA and TST administration. Fifteen participants were lost to follow-up, 6 were unreachable, 7 did not keep appointments and 2 refused consent (figure 1). Of these 76 participants, 62 tested IGRA negative on second testing, thirteen tested IGRA positive and there was one indeterminate result.

From the 62 who tested IGRA negative with the second test, four participants did not return for the TST reading and a third IGRA. Of the 58 participants who returned for a third visit (3 days later), three had a positive third IGRA result (2 TST = 0mm, 1 TST = 15mm) and four did not have a third IGRA (TST = 7, 7, 10, 16mm). In total 51 participants tested IGRA negative for the third time, 48 had TST readings of 0 mm and two had TST readings of 1 mm. Only 1 participant had a TST reading of 6 mm (figure 1). The 48 participants with persistent IGRA negative and TST readings of 0mm were defined as HITTIN.

One of the thirteen participants with a positive second IGRA result was excluded due to presenting with TB symptoms. The remaining twelve participants had the following IGRA results and TST readings (figure 1). Two participants had a third IGRA positive result (TST 10, 20mm), two did not have a third IGRA (TST 13, 19mm) and eight had negative third IGRA result (6 TST = 0mm, 1 TST = 1mm, 1 TST = 20mm). There were six participants who converted from IGRA negative to positive with the second IGRA, tested negative with the third IGRA and had TST readings of 0 mm. Upon closer investigation of these individuals we noticed that 4 of the positive results on the second IGRA were close to the cut-offs determined by the manufacturer’s algorithm (see figure 2 A and B). Three participants who tested negative on the first and on the second IGRA tested positive on the third IGRA done 3 days after TST administration. Although the three IGRA positives were likely due to boosting, we did not include them in the HITTIN subject count.[14]

**Figure 2:**
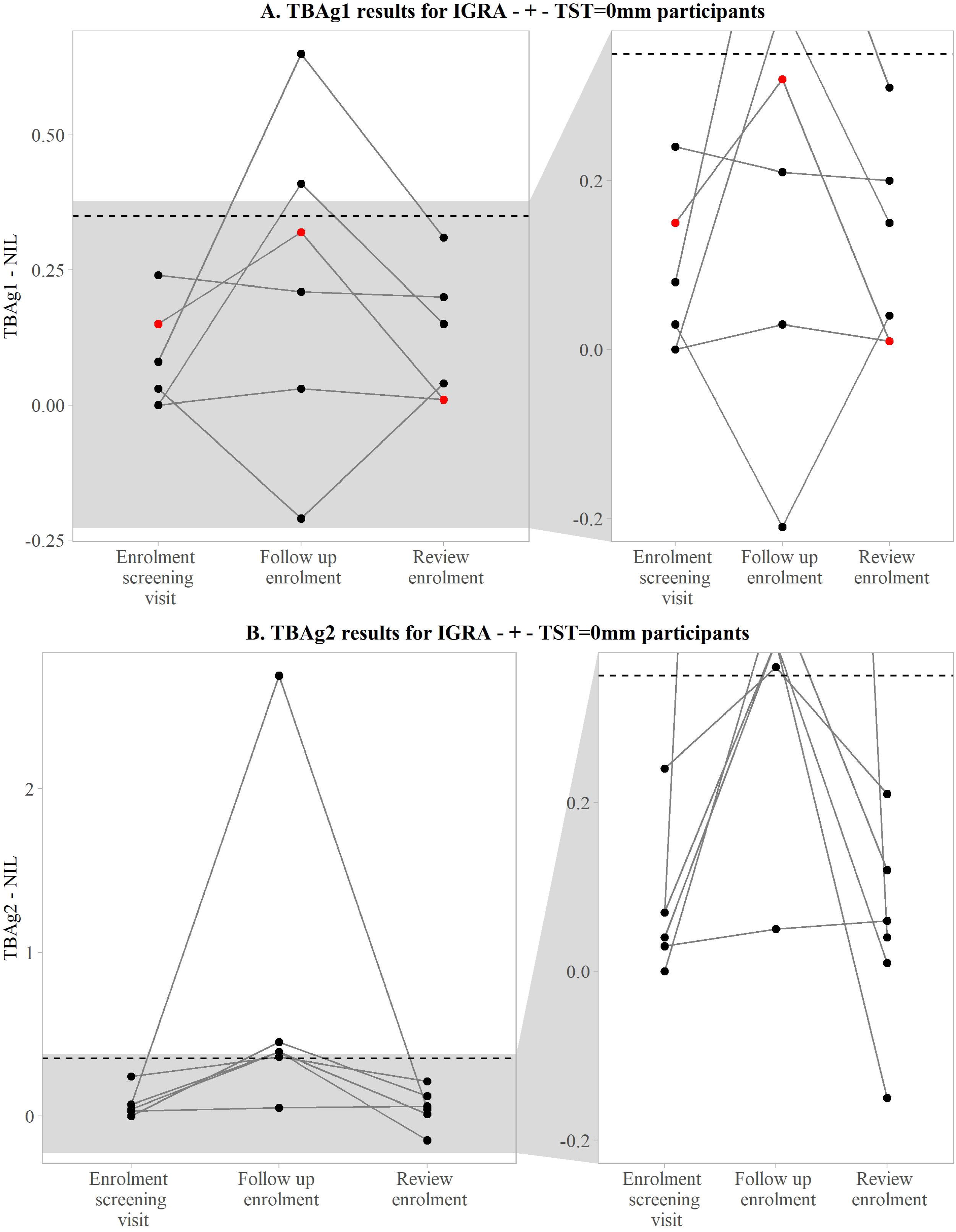
TBAg1 and TBAg2 responses. A) TBAg1^a^ and B) TBAg2^b^ IGRA^c^ results of the six participants who converted during visit 2 (follow up enrolment) and reverted after 3 days (review enrolment). Each of the six participants had TST^d^ readings of 0 mm. The threshold of 0·35 IU/ml is indicated by the dashed line. ^a^ TBAg1 (TB1 tube result for QuantiFERON®-TB Gold Plus test) ^b^ TBAg2 (TB2 tube result for QuantiFERON®-TB Gold Plus test) ^c^ IGRA (interferon gamma release assay) ^d^ TST (tuberculin skin test)

We repeated the TST using PPD-RT23 on average 402 days after the first administration of PPD-S in 8 of the participants (table 1). The results were identical for TST=0mm (6 subjects) or near identical for TST > 5 mm (18mm vs 16mm and 28mm vs 26mm on the first read) which is consistent with previous reports.[31] Five of these six participants are IGRA negative and one tested IGRA positive.

**Table 1:** Comparison of TST after PPD-S and PPD-RT23 administration.

| **Participant number** | **IGRA<sup>a</sup>** | **Previous TST<sup>b</sup> reading in mm (PPD<sup>c</sup>-S)** | **Previous TST date** | **Repeat TST reading in mm (PPD-RT23)** | **Repeat TST reading date** |
| --- | --- | --- | --- | --- | --- |
| **2RTB0013** | Positive | 16 | 30/01/2018 | 18 | 21/01/2019 |
| **2RTB0186** | Negative | 0 | 17/11/2017 | 0 | 21/01/2019 |
| **2RTB0253** | Negative | 0 | 23/11/2017 | 0 | 21/01/2019 |
| **2RTB0165** | Negative | 0 | 16/11/2017 | 0 | 24/01/2019 |
| **2RTB0183** | Negative | 0 | 17/11/2017 | 0 | 24/01/2019 |
| **2RTB0135** | Negative | 0 | 01/12/2017 | 0 | 24/01/2019 |
| **2RTB0015** | Positive | 0 | 30/01/2018 | 0 | 25/01/2019 |
| **2RTB0005** | Positive | 25 | 02/02/2018 | 28 | 28/01/2019 |

<sup>a</sup> IGRA (interferon gamma release assay)
<sup>b</sup> TST (tuberculin skin test)
<sup>c</sup> PPD (purified protein derivative)

Demographic and clinical features of the study participants are listed in Table 2. Age and sex were not significantly different between the HITTIN, HIT, and participants who had previous TB. HITTIN were 85·4% female, with an average age of 43·3±5·6 years. The HIT group was 85·7% female with an average age of 42·9±5·6 years, and participants who had previous TB were 69·2% female and had an average age of 43·0±6·9 years. HITTIN were diagnosed with HIV on average 8·0±3·0 years before enrolment. This differed slightly but significantly from the HIT group (p=0·01) who were diagnosed with HIV on average 10·0±3·8 years before enrolment and from the previous TB group (diagnosed 99±3·1 years before enrolment, p=001, unpaired t test). There was also a significant difference (p=0·04, unpaired t test) between time spent on ART before enrolment in that HITTIN were on ART for a slightly shorter time (7·0±3·0 years), compared to the HIT group (8·0±3·0 years) and previous TB group (8·5±2·2 years on ART, p=001). There was however no significant difference between time from HIV-1 diagnosis to ART initiation between HITTIN (10±2·0) and the HIT group (2·0±3·0, p=0·09, unpaired t test), nor between HITTIN and the previous TB group (1-4±2-3, p=031, unpaired t test). A total of 5/48 (10%) of HITTIN were not virally suppressed (V^100 copies) compared to 0/35 HIT. There was no statistical difference (p=0·07, Fisher’s exact test). The average time between the last CD4 count and the first IGRA test for HITTIN (2·3±2·0 years) was significantly less than the HIT group (8-7±3-4 years, p < 0·00001, unpaired t test), but did not differ significantly from the previous TB group (Arm2) (2·7±21 years, p = 0·33, unpaired t test). Participants did not report any Stage 3 or 4 HIV related illnesses in the period between these two events. HITTIN (79·2%) were not more likely to have received Isoniazid preventive therapy (IPT) compared to the HIT group (914%) (p=022, Fisher’s exact test). There were no other significant epidemiological or demographic factors that differed between the HIT and the HITTIN group. HITTIN had significantly higher BMI levels (28·2±6·2) compared to participants with previous TB (25·5±6·0, p=0·04, unpaired t test), but not compared with the HIT group (29·2±5·9, p=0·46, unpaired t test). The average time between the first and second IGRA for HITTIN was 203±151 days and for the HIT group 292±70 days (p=0002, unpaired t test).

**Table 2:** Demographic data.

|  | HITTIN<sup>a</sup> | (%) | HIT<sup>b</sup><br>(IGRA double+,<br>TST>5mm) | (%) | p-value<sup>d</sup> | HIV-1-infected,<br>Previous TB<sup>c</sup> | (%) | p-value<sup>d</sup> |
| --- | --- | --- | --- | --- | --- | --- | --- | --- |
| **n** | 48 |  | 35 |  |  | 39 |  |  |
| **Age at enrollment** | 43.3±5.6 |  | 42.9±5.6 |  | 0.72 | 43.0±6.9 |  | 0.79 |
| **Sex (% female)** | 41 | 85.4 | 30 | 85.7 | >0.99 | 27 | 69.2 | 0.12 |
| **Income range** |  |  |  |  |  |  |  |  |
| R0-R3000 | 27 | 56.3 | 22 | 62.8 |  | 29 | 74.4 |  |
| R3001-R5000 | 16 | 33.3 | 8 | 22.9 |  | 5 | 12.8 |  |
| Above R5000 | 5 | 10.4 | 5 | 14.3 | 0.56 | 5 | 12.8 | 0.26 |
| **Education level** |  |  |  |  |  |  |  |  |
| No schooling | 0 | 0.0 | 0 | 0.0 |  | 1 | 2.6 |  |
| Primary school | 3 | 6.3 | 7 | 20.0 |  | 3 | 7.7 |  |
| High school | 41 | 85.4 | 28 | 80.0 |  | 35 | 89.7 |  |
| Diploma/Degree | 4 | 8.3 | 0 | 0.0 | 0.09 | 0 | 0.0 | 0.69 |
| **Employed** | 26 | 54.2 | 19 | 54.3 | >0.99 | 17 | 48.9 | 0.39 |
| **Weight (kg)** | 78.1±17.7 |  | 82.7±17.8 |  | 0.27 | 72.2±17.5 |  | 0.11 |
| **Height (m)** | 1.7±0.1 |  | 1.7±0.1 |  | 0.29 | 1.7±0.1 |  | 0.17 |
| **BMI<sup>e</sup>** | 28.2±6.2 |  | 29.2±5.9 |  | 0.46 | 25.5±6.0 |  | 0.04 |
| **Years since HIV<sup>f</sup> diagnosis (from diagnosis to 1st enrolment)** | 8.0±3.0 |  | 10.0±4.0 |  | 0.01 | 9.9±3.1 |  | 0.01 |
| **Time between HIV diagnosis and TB<sup>g</sup> diagnosis** | Not applicable |  | Not applicable |  |  | 1.8±2.9 |  |  |
| **Age at TB diagnosis** | Not applicable |  | Not applicable |  |  | 35.0±7.5 |  |  |
| **Time on ART<sup>h</sup> in years (from ART start to 1st enrolment)** | 7.0±3.0 |  | 8.0±3.0 |  | 0.04 | 8.5±2.2 |  | 0.01 |
| **Time in years since HIV diagnosis and ART start** | 1.0±2.0 |  | 2.0±3.0 |  | 0.09 | 1.4±2.3 |  | 0.31 |
| **CD4 count before ART initiation** |  |  |  |  |  |  |  |  |
| < 200 CD4+ cells/mm3 | 47 | 97.9 | 33 | 94.3 |  | 36 | 92.3 |  |
| Between 200-350 CD4+ cells/mm3 | 1 | 2.1 | 2 | 5.7 | 0.57 | 3 | 7.7 | 0.32 |
| **Most recent CD4 count** | 505.8±191.4 |  | 486.9±193.8 |  | 0.66 | 545.6±248.6 |  | 0.40 |
| **Last viral load (no VL done at diagnosis)** |  |  |  |  |  |  |  |  |
| LDL<sup>i</sup> | 27 | 56.3 | 20 | 57.1 |  | 23 | 59.0 |  |
| < 20 | 5 | 10.4 | 9 | 25.7 |  | 4 | 10.3 |  |
| <100 | 11 | 22.9 | 6 | 17.1 |  | 3 | 7.7 |  |
| 100-1000 | 4 | 8.3 | 0 | 0.0 |  | 7 | 17.9 |  |
| >1000 | 1 | 2.1 | 0 | 0.0 | 0.09 | 2 | 5.1 | 0.82 |
| **AIDS defining illness in last year** | 0 | 0.0 | 0 | 0.0 |  | 0 |  |  |
| **IPT<sup>j</sup> received** | 38 | 79.2 | 32 | 91.4 | 0.22 | 38 | 97.4 | 0.02 |
| **Current IPT** | 5 | 10.4 | 1 | 2.9 | 0.39 | 4 | 10.3 | 1.00 |
| **Cotrimoxazole used** | 24 | 50.0 | 23 | 65.7 | 0.18 | 23 | 59.0 | 0.52 |
| **Previous interventional studies** | 0 | 0.0 | 0 | 0.0 |  | 0 | 0.0 |  |
| **Other illness** |  |  |  |  |  |  |  |  |
| Diabetes | 0 | 0.0 | 1 | 2.9 |  | 1 | 2.5 |  |
| Asthma | 1 | 2.1 | 0 | 0.0 |  | 0 | 0.0 |  |
| Hypertension | 13 | 27.1 | 6 | 17.1 |  | 4 | 10.3 |  |
| Other | 2 | 4.2 | 0 | 0.0 | 0.22 | 0 | 0.0 | 0.02 |
| **Recreational substances (cannabis)** | 3 | 6.3 | 1 | 2.9 | 0.63 | 2 | 5.1 | 1.00 |
| **Alcohol use** |  |  |  |  |  |  |  |  |
| No alcohol | 33 | 68.8 | 28 | 80.0 |  | 27 | 69.2 |  |
| Occasionally | 15 | 31.2 | 7 | 20.0 | 0.32 | 12 | 30.8 | 1.00 |
| **Smoking (Yes)** |  |  |  |  |  |  |  |  |
| Current | 2 | 4.2 | 1 | 2.9 |  | 4 | 10.3 |  |
| Previous | 0 | 0.0 | 0 | 0.0 | >0.99 | 2 | 5.1 | 0.13 |

<sup>a</sup> HITIN (HIV-1-infected persistently TB, tuberculin and IGRA<sup>k</sup> negative)
<sup>b</sup> HIT (HIV-1-infected IGRA positive tuberculin positive)
<sup>c</sup> HIV-1-infected, Previous TB: Subjects with a history of bacteriologically confirmed TB during ART that had been cured for at least 3 years (Arm 2)
<sup>d</sup> The initial p-value refers to the comparisons between HITIN and the HIT group. The second p-value here refer to the comparisons between HITIN and previous TB group. Categorical frequencies between the HITIN and HIT groups were compared using the Fisher's exact or $\chi^2$ test. Means were compared using the unpaired t test.
<sup>e</sup> BMI (body mass index) was calculated as weight (kg)/height<sup>2</sup> (m).
<sup>f</sup> HIV (human immunodeficiency viruses)
<sup>g</sup> TB (Tuberculosis)
<sup>h</sup> ART (Antiretroviral therapy)
<sup>i</sup> LDL (lower than detectable)
<sup>j</sup> IPT (Isoniazid preventive therapy)
<sup>k</sup> IGRA (interferon gamma release assay)

To determine if HITTIN had prior contact with *Mtb*, serum IgG titres were measured by ELISA against Mtb-specific antigens ESAT-6 and CFP-10 (Figure 3). Initially only a subset of n=15 was selected for the ELISA as the work was exploratory. In the subset of HITTIN tested, approximately 73·3% (11/15) and 91·7% (11/12) of participants with active TB had evidence of antibody responses to ESAT-6/CFP-10. To further decipher the differences between HITTIN and HIT across various TB-associated antigens and isotypes beyond general *Mtb* exposure, a customized Luminex assay was performed (Figure 4A). The analysis included the samples used in the initial ELISA. The Luminex data replicated and expanded upon the ESAT-6/CFP-10 ELISA data and similar levels of IgG, IgA1 and IgM for ESAT6/CFP10, PPD and LAM between HITTIN and HIT were observed. While there were no significant differences between the two groups, there was a broad spectrum of responses, especially for IgG and IgA2 for ESAT-6/CFP-10 and PPD and IgM LAM within the groups. This indicated a heterogenic response across individuals in the absence of association with the HITTIN phenotype, suggesting that both groups were indeed exposed and potentially infected.

**Figure 3:**
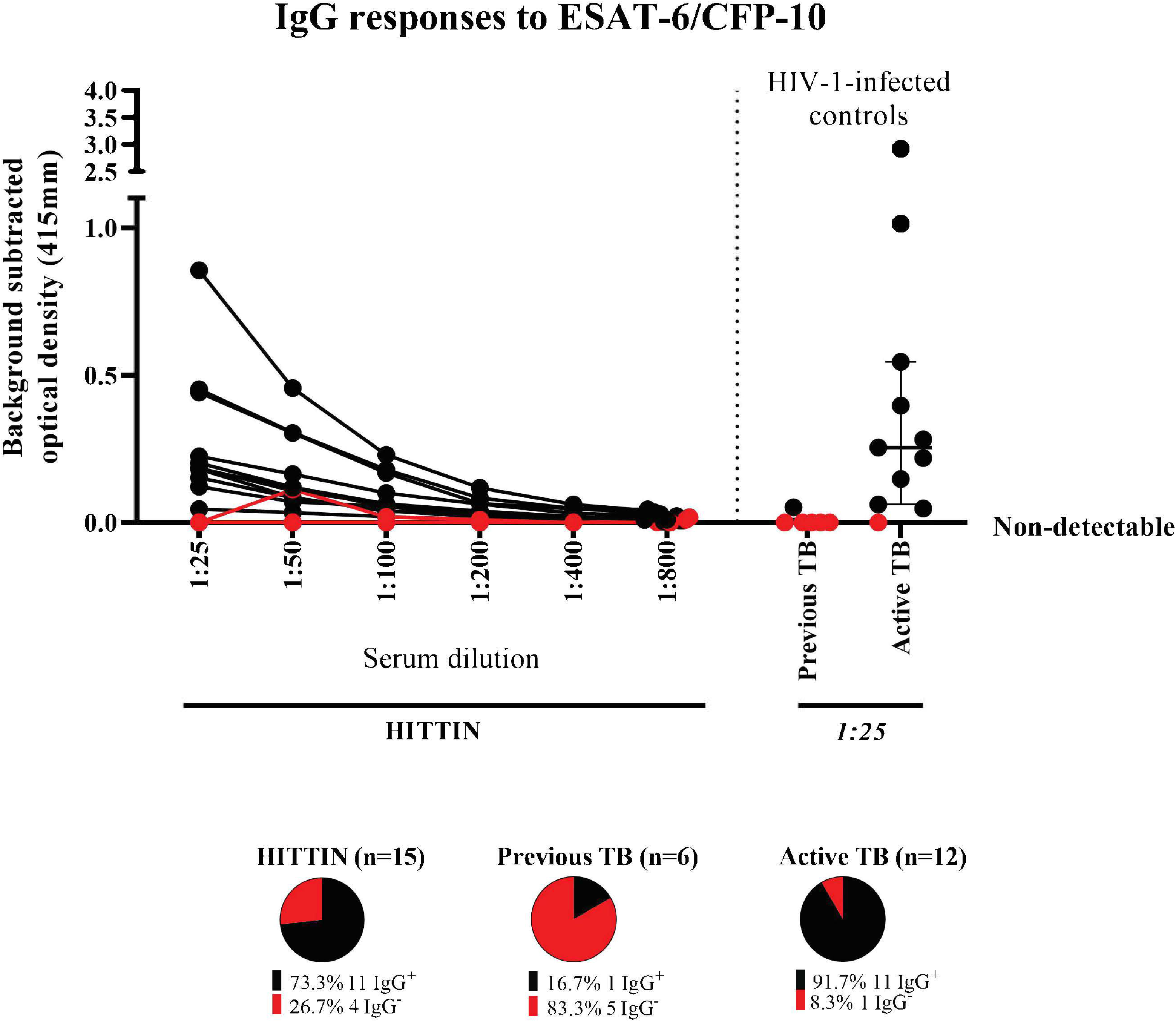
IgG responses to ESAT-6/CFP-10. IgG antibody titers against *Mtb*specific antigens ESAT-6/CFP-10 (4μg/ml). Titration curves of serum antibody responses against a fusion protein of ESAT-6 and CFP-10 (Lionex) in 19 HIV-1-infected persistently TB, tuberculin and IGRA negative (HITTIN). Optical density (OD 415mm) values from antigen-uncoated wells at each dilution were subtracted from serum OD values at that dilution. A positive response was determined by OD values above background, and absent binding curves are demonstrated in red. Controls include 6 individuals with previous TB from Arm 2, and HIV-1-infected controls with active TB with median and interquartile ranges shown. Out of 15 HITTIN tested, 13 demonstrated decreased OD on dilution (68·4%), whereas only 1/6 (16·7%) of Arm 2 participants did (16·7%), and 11/12 (91·7%) controls with active TB. There is no statistically significant difference between IgG+ antibodies in HITTIN compared to the previous TB (Arm 2) group (p=0·18, Kruskal-Wallis test with Dunn’s post test correction for multiple comparisons) and active TB group (p=0·06, Kruskal-Wallis test with Dunn’s post test correction for multiple comparisons).

To further investigate the overall antibody profile of the two groups in a more comprehensive way, a supervised multivariate analysis was applied. Therefore, a partial least-square discriminant analysis (PLSDA) using a least absolute shrinkage and selection operator (LASSO) was performed. The PLSDA loadings plot (Figure 4C) showed the selected minimal features accounting for the maximal variance in the data set. Features enriched in HIT are coloured in blue and trend towards the right side of the plot, whereas features higher in HITTIN (green) are on the left side of the plot. However, little separation can be observed in the PLSDA scoring plot (Figure 4B), indicating that the antibody profiles between HITTIN and HIT are similar and the groups cannot be separated based on their antibody features.

**Figure 4:**
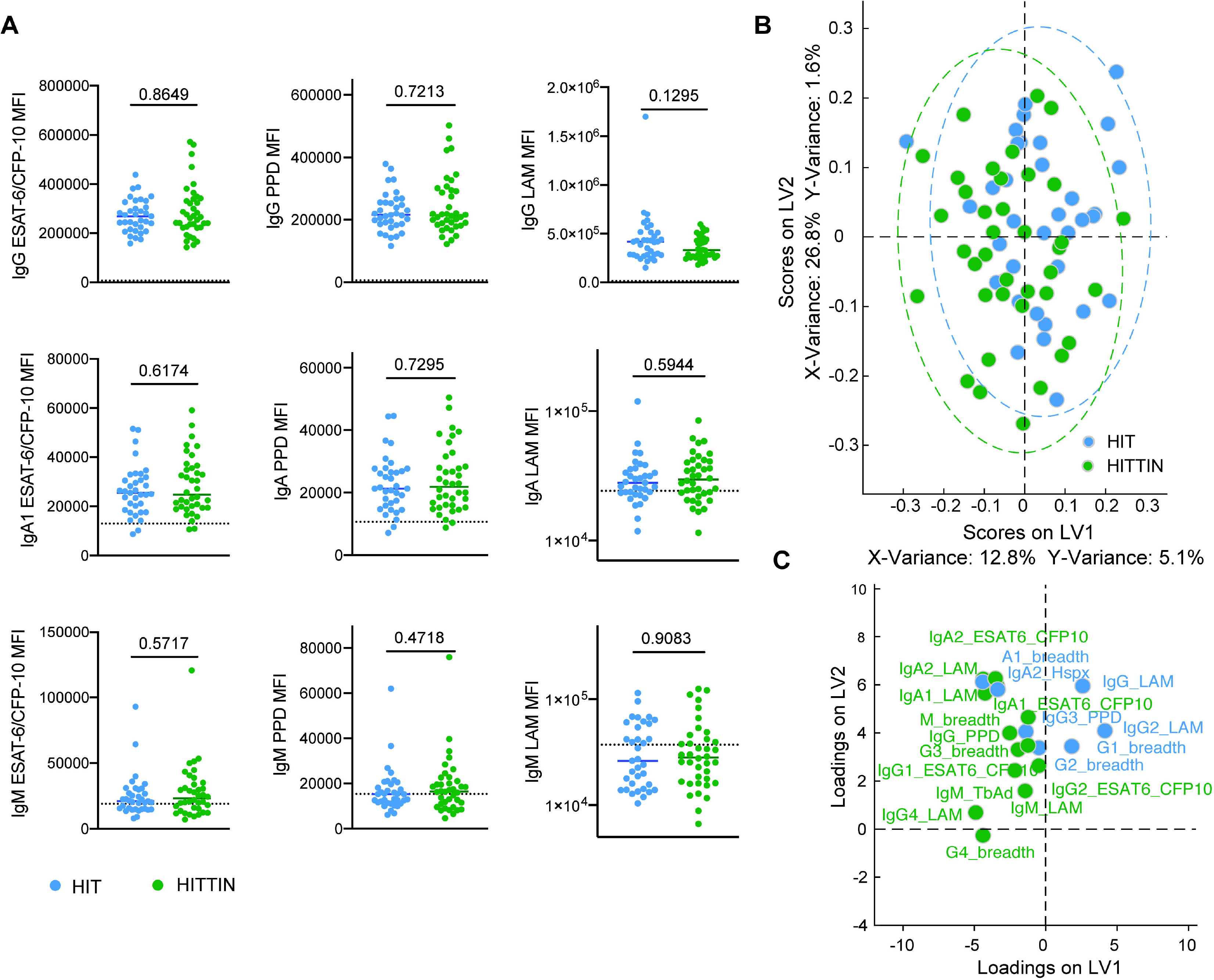
HITTIN and HIT individuals harbor similar antibody profiles across subclasses/isotypes and antigens. **A:** Plasma levels of IgG, IgA1 and IgM reactive to ESAT6/CFP10, PPD and LAM were profiled across HIV-1- infected persistently TB, tuberculin and IGRA negative (HITTIN) (n=38) and HIV-1-infected IGRA positive tuberculin positive (HIT) individuals (n=35) represented by median fluorescent intensity (MFI) using a customized Luminex assay. The antigen-specific isotype levels are plotted for the two different groups, HITTIN (green) and HIT individuals (blue), the median is depicted, the dotted line represents the median level detected in HIV-negative, healthy North American volunteers. Mann-Whitney test was used to compare between groups. **B and C:** LASSO- PLSDA was performed utilizing antigen-specific antibody levels (total number of initial features=48). The scores plot (**B**) shows the distribution of groups (each dot represents one individual) across two latent variables (LV1 and 2), driven by the down selected features (**C**). P-values shown in *figure 4* calculated with Mann-Whitney test.

## Discussion

We present the description of a stringently defined phenotype of persons without previous TB who test persistently negative to IGRA and tuberculin within a highly TB susceptible HIV+ population in the Western Cape, where *Mtb* transmission is endemic in the community.[18] We targeted individuals with prolonged and persistent *Mtb* infection exposure using age as a surrogate for exposure frequency. Given that a substantial proportion of subjects enrolled in our study display the HITTIN phenotype (48/286) this highlights the importance of the phenotype for TB control and a proper understanding of the flow of *Mtb* through exposed populations.

A unique aspect of our study was the enrolment of HIV+ persons who had undergone a period of low CD4 counts. There is ample epidemiological evidence that risk of TB increases greatly for such patients. The HITTIN cases defined in our study show a durable TB resistance phenotype that may be independent of IFN-γ cell mediated and delayed-type hypersensitivity (DTH) immune responses. They remained free of TB disease and tested TST and IGRA negative many years (80±3·0) after HIV diagnosis. Importantly, even HIV+ persons receiving ART such as our participants, and therefore considered immune reconstituted, still have at least double the risk of acquiring active TB compared to the general population. [34]

Immune reconstitution was key since we based the HITTIN phenotype on immune assays which are dependent on functioning T-cells. The ability of ART reconstituted CD4 T-cells to produce IFN-γ in response to ESAT-6/CFP-10 remains the same or improves with time on ART, although this is not always fully restored to the same extent as in HIV uninfected persons.[35-38] Although the ART duration in HITTIN was shorter than that of HIT and previous TB groups, it has been determined that the functional potential of *Mtb*-antigen specific T-cells can significantly change after only three months on ART.[36] Most studies considered the effect of ART over only a few weeks, but HITTIN identified in this study have been on ART an average of 7 years, making it unlikely that the negative results in IGRA and TST reported here are due to a lack of host ability to produce IFN-γ.

In the study area, TST responses of ≥10 mm are considered indicative of immune conversion and infection. Multiple studies have shown that TST readings of ≥10 mm have very low reversion rates (<10%). Given that such TST responses are very stable, it is highly unlikely that our negative TST measurements =0 mm reflected previous positive readings which reverted but were most likely persistently negative including during the period of low CD4+ T-cell counts. [39-42] Nevertheless, negative TST or IGRA results could be the result of a lack of sufficient exposure despite being in a high incidence environment. This seemed unlikely, since studies have shown that *Mtb* transmission occurs very frequently within this community.[21] More definite, however, the presence of IgG ESAT- 6/CFP-10 antibodies in our study indicated previous exposure to *Mtb* of these individuals as previous vaccination with BCG would not have accounted for the presence of these antibodies.[43] While multiple antigens from *Mtb* have been tested here, including the *Mtb* specific ESAT6/CFP10 antigens, the possibility of detecting nontuberculosis mycobacteria (NTM) cross-reactive antibodies exists. ESAT-6/CFP-10 are RD1 encoded proteins and are used in the QFT assay due to well-known deletion in BCG. The same organisms producing ESAT-6 homologues (e.g. *Mycobacterium marinum* and *Mycobacterium kansassi)* can give a falsely reactive IGRA result [44,45]. A role of NTM-specific antibody detection should be taken into consideration when evaluating the results and deserves more careful study.

Even though HITTIN cases were diagnosed with HIV for a shorter time than the HIT group and previous TB participants before enrolment, one cannot infer that this implies that they were infected with HIV for a significantly shorter time, as time from HIV infection is impossible to determine. Importantly, there was no significant difference between time since HIV diagnosis and initiation of ART. The average time between the last CD4 count and the first IGRA was significantly shorter in HITTIN compared to HIT, which minimizes the risk of false negative immune conversion results. There were no other significant differences between HITTIN and the HIT control group.

The initial ELISA tests displayed in figure 3 show absence of IgG recognition of ESAT-6/CFP-10 in % (4/15) of the HITTIN. This by itself does not necessarily infer lack of prior infection with *Mtb*, as not all HIV+ controls with microbiologically confirmed active TB (~90%) had antibodies against ESAT-6/CFP-10 and is in keeping with prior literature on heterogeneity in the human antibody response against *Mtb*.[46] The similarity in antibody levels across HITTIN and HIT individuals was confirmed across different antigens, including PPD, ESAT-6/CFP-10 and LAM and upon different antibody isotypes shown in figure 4. The ESAT-6/CFP-10 results were cross-validated by two assays. The existence of class-switched IgG and IgA antibodies suggested IFN-γ-independent CD4 T-cell help despite negative IGRA and TST testing, supporting the claim of *Mtb* exposure in HITTIN.[12] The lack of separation between HIT and HITTIN in the PLSDA (Figure 4B and C) confirmed the similar antibody profiles observed in these groups in HIV+ persons, concurrent with previous findings in an HIV-uninfected cohort in Uganda. [12] The presence of specific *Mtb*-antibodies suggests that HITTIN cases may have been transiently infected with *Mtb* and yet do not display *Mtb*-specific IFN-γ production and TST reactivity.

It is difficult to elucidate whether HITTIN have cleared infection or are still currently infected. It might be that infection was cleared after *Mtb* exposure and that T-cell differentiation was halted prior to the development of IFN-γ specific Th1 cells, thus favouring early clearance rather than a resistance to *Mtb*-specific IFN-γ immunoconversion phenotype. [12] This could account for the lack in *Mtb*-specific IFN-γ production but not the lack of TST reactivity. The HITTIN phenotype is therefore strengthened by combining IGRA and TST negativity.

Similarly to HITTIN, the HIT group likely captures a spectrum of *Mtb* infection. Positive TST and IGRA readings could indicate present or cleared infection. Without a definitive microbiological standard it is difficult to differentiate between the two. Persons treated for previous TB could remain TST positive for up to nine years after treatment.[39,40] Also, not all persons with positive TST develop TB when they become immunosuppressed, including persons who are HIV+.[39,47-50] Importantly, the HIT group also has no history of previous or current TB and could therefore also contain an interesting subgroup of persons who resist progression to TB. Interestingly, another cohort recruited in Worcester, South Africa found an increased TB incidence rate for those with a baseline positive TST and IGRA (0.6 cases per 100 person years [95% CI 0.43-0.82], 0.64 cases per 100 person years [0.45–0.87]), compared to participants who had baseline negative TST and IGRA results (0.22 cases per 100 person years (0.11-0.39), 0.22 cases per 100 person years (0.12-0.38).[51] In contrast, presently we do not know the significance of antibody positivity and if this positivity entails increased or reduced risk of TB disease. This question can indeed only be answered by long term follow-up of antibody positive and negative subjects.

Our study is enriched for female (214/268 Arm 1 and 27/39 Arm 2) compared to male participants and 41/48 (85·4%) of the identified resisters are female. In South Africa the HIV prevalence is higher in woman (20·6%), compared to men (148%).[19] In addition to this men are also less likely to attend clinics and be tested for HIV and typically have lower ART coverage.[52,53] This study is biased in that it is directed to identify a highly selected phenotype. We identified participants from Adherence Clubs at HIV clinics in the community which biases selection towards HIV+ persons who are healthy and ART adherent. Members of the ART club are perhaps more likely to make healthier social choices and elect not to smoke or consume excessive amounts of alcohol as was found in our study. A limitation of community-based *Mtb* exposure studies is the inability to define and measure the exact extent of exposure. We assume the exposure is consistent among persons residing in these high exposure environments and base the assumption of exposure on research showing that exposure occurs mostly within the community rather than the household. [18] Lastly we excluded previous TB based on patient and clinical record history and not based on chest x-ray (CXR). Studies show that both questionnaires and CXRs have low negative predictive values to identify previous TB [54]. Medical folders were screened and evaluated for each enrolled patient. Future studies include BALs for a subset of the identified HITTIN group, as well as TB GeneXpert testing and CXR.

The 48 identified HITTIN (17%) is likely an underestimate based on the likelihood of false positives identified due to characteristics of the assay. The likely 6 false positive participants had initially negative IGRA results (see figure 1 and 2). On retesting and just prior to TST administration, these results converted to positive and three days later, they reverted to negative with TST = 0mm. This is most likely a false positive IGRA result on the second visit. If we assume a similar misclassification rate at the first enrolment IGRA test, the count of HITTIN might have increased considerably. We also did not include participants who had three consecutive negative IGRA results with TST readings of 1mm. This is a minimal reaction, if any, and should these participants be included in addition to the false positives, there were a potential 56/286 (20%) HITTIN. In either case we think that 17% HITTIN is a low end estimate further emphasizing the importance of this phenotype in *Mtb* transmission.

This is the first description of HIV+ persons who remain free of TB and test persistently negative by IGRA and TST. We identified a unique group of individuals on longterm ART, in Cape Town, a high TB incidence environment. Despite having experienced very low CD4 counts and high exposure to *Mtb*, confirmed by the presence of antibodies, these HITTIN remained persistently IGRA and TST negative and did not develop TB before or after HIV diagnosis. Although the nearly 20% of HITTIN identified are not representative of the entire HIV population, our study highlights that this phenotype can be identified amongst HIV+ persons and represents a valuable phenotype. Even though HITTIN is a stringently defined and unique phenotype it is a critical research focus for prevention of TB in HIV-infected persons.

## Data Availability

The data that support the findings of this study are available from the corresponding author, Dr. Möller, upon reasonable request.

## Contributors

ES, EH, MM, CK, RJW, MO, GW and EEK conceptualised the study and the phenotype. SB, MM and EEK designed the database. EEK and MM were involved with participant recruitment and enrolment. ES, MM, EH, SB, CK, and EEK coordinated the project and SB, MM, CK and EEK were also involved with project administration. SB established the laboratory, procured laboratory equipment in Cape Town, South Africa and MO in Montreal, Canada. SB processed and analysed IGRA samples. AJ, SF, SS and GA did the antibody work and data analysis. EEK, ES, MM, AJ and SF drafted the manuscript and all co-authors reviewed and edited the final manuscript. All authors approved the final manuscript as submitted and agree to be accountable for all aspects of the work.

## Acknowledgements

We would like to acknowledge and thank the following individuals for their contribution to the ResisTB study: Johannes Taljaard, Mark Cotton, Hans Prozesky, Marije van Schalkwyk, Stefanus Malherbe, Elizna Maasdorp, Gian van der Spuy, Anna Coussens, Allison Seeger, Molebogeng Rangaka, Grant Theron, Suventha Moodley, Nosipho Mtala, Siyabulela Justice Mboniswa, Janice Theys, Glenda Durrheim, Naomi Okugbeni and Vuyiswa Ndunana.

The study was approved by the Health Research Ethics Committee (HREC) of Stellenbosch University [N16/03/033 and N16/03/033A] and the Faculty of Health Sciences Human Research Ethics Committee of the University of Cape Town [HREC 755/2016 and 702/2017]. Additional approval was obtained from the City of Cape Town and Western Cape government for access to the relevant clinics.

The work herein was made possible through funding by the South African Medical Research Council through its Division of Research Capacity Development under the SAMRC Clinician Researcher M.D PhD Development programme. The content of any Publications from any studies during this Degree are solely the responsibility of the authors and do not necessarily represent the official views of the South African Medical Research Council. This publication is supported by NeutroTB which is part of the EDCTP2 programme supported by the European Union (grant number TMA2018CDF-2353-NeutroTB). The views and opinions of authors expressed herein do not necessarily state or reflect those of EDCTP.

## Data Sharing Statement

Data is available on reasonable request to the corresponding author.

